# Epidemic Dynamics and Non-Pharmaceutical Interventions: A Mathematical Approach

**DOI:** 10.1101/2025.06.08.25329232

**Authors:** Davi Vieira Ramos de Oliveira, Marina Lima, Graciele Paraguaia Silveira, João Frederico da Costa Azevedo Meyer

## Abstract

We present a comparative analysis of three compartmental mathematical models aimed at capturing the dynamics of directly transmitted infectious diseases while explicitly incorporating social behaviour and non-pharmaceutical interventions (NPIs). The models are extensions of the classical SEIR framework and introduce heterogeneous social behaviours, behaviour switching mechanisms over time, and vaccination strategies. In Model 1, we consider a closed population with two subgroups differing in their adherence to NPIs. Model 2 incorporates vital dynamics and a hospitalization compartment, allowing long-term analysis. Model 3 further extends the framework by including vaccination and dynamic behavioural changes driven by the evolution of the epidemic. Through numerical simulations, we assess the impact of behavioural heterogeneity, timing of vaccination, and post-recovery behaviour on epidemic curves. The results show that early adoption of NPIs and timely vaccination significantly reduce infection peaks and cumulative cases. Behavioural transitions, if not sustained, can lead to secondary outbreaks. Our findings reinforce the importance of integrating behavioural and social aspects into epidemiological models to better inform public health policies and intervention planning.

## Introduction

Directly transmitted diseases represent a significant challenge for public health systems, given their ability to spread rapidly among susceptible individuals and cause major social and economic impacts. Over the last few decades, mathematical models of epidemics have played a fundamental role in understanding the spread of these diseases, with a SIR-type modelling, as proposed by Kermack and McKendrick [1], this model as well as its many variations being one of the most influential type of models in mathematical epidemiological modelling. However, the inclusion of behavioural factors in such models has become increasingly important, especially in recent contexts, such as the COVID-19 pandemic [2].

Social behaviours, such as social distancing, wearing masks, and quarantine decisions, for example, directly affect transmission dynamics, altering contact rates between individuals and the progression of the disease in a population. Studies such as those by [3] and [4] have shown that the incorporation of behavioural interventions can substantially modify the predictions of epidemic outbreaks and the effectiveness of control strategies. Thus, in agreement with these authors, there is a need for models that not only capture the biology of transmission, but also integrate behavioural and social aspects, allowing for a more realistic analysis of the impact of non-pharmacological interventions.

Mathematical models are fundamental tools for analysing the dynamics of disease transmission, allowing the identification of contagion patterns, risk factors, and vulnerable populations. However, traditional models face significant limitations, such as the assumption of population homogeneity and the simplification of contact dynamics. These restrictions can compromise the accuracy of the results and the applicability of possible predictions. In order to improve the effectiveness of these models, it has become increasingly necessary to incorporate more realistic interactions that take into account population heterogeneity and the adaptive behaviour of individuals.

In this paper, we propose a comparative analysis of three mathematical models that incorporate different aspects of social behaviours into the dynamics of diseases transmitted directly. The models presented are also variants of classical compartmental models, adjusted to include parameters that reflect changes in social behaviours of a population during epidemic outbreaks. Specifically, we introduced the parameter ‘*ρ*’, which represents the probability of a recovered individual returning to the susceptible state without social distancing, and the parameter ‘*ϑ*’, which captures the change in behaviour over time caused by the evolution of the epidemic, as well as economic and social pressures.

## Mathematical Modelling and Simulations

The purpose of this section is to introduce epidemiological developments of SEIR-type mathematical models for describing and analysing the spread of infectious diseases in specific populations, as well as estimating the speed of the disease’s dissemination in the presence of behavioural changes. Thus, these epidemiological models will be discussed with their variations, considering aspects such as the presence or absence of vital dynamics and vaccination efforts. Furthermore, we will include new aspects in the classical models, such as division of the population into different social behaviours, the probability of changes in such behaviours after a recovery from the disease and changes in attitude towards mitigation measures during the epidemic outbreak.

The compartmental models proposed have been (and still are) widely discussed in the literature [5–11]. In spite of there being several studies that use both classical models and those that incorporate social behaviours, we did not find references in the literature that consider the division of the population into two types of social behaviours and behavioural changes during the outbreak or after individuals’ recovery from the disease, as discussed in this work.

As mentioned above and based on adaptations of the classic SEIR model, we propose three compartmental models, all of them of Kermack and McKendrick type, for investigating the spread of directly transmitted infectious diseases. These models aim to be applicable to a wide range of diseases with this mode of transmission. For the study, the population is divided into those who do not use non-pharmacological strategies to prevent spread of a disease and another group who adhere to them. Susceptible individuals who do not use non-pharmacological strategies are at greater risk of being exposed and infected, whereas those who do use these strategies have a lower probability of becoming infected. These non-pharmacological interventions, such as use of masks as well as social distancing, play an important role in slowing the spread of a disease, allowing time for the development and distribution of vaccines and drugs. In this text, we will use the expression “social distancing” to identify not only isolation choices but other non-pharmaceutical mitigation strategies. Social distancing (in the sense indicated above) has a non-linear impact on the effectiveness of vaccination, highlighting the importance of maintaining safe behaviours even with the availability of vaccines [12]. The proposed models in this work therefore allow us to evaluate how the adoption of non-pharmacological strategies and changes in social behaviour can impact the spread of the disease in a given population.

In all three models, we consider that the population is divided into two subgroups with different behaviours regarding social distancing during the beginning of a disease outbreak. Using variations of the classic SEIR model, we denote by *S*_*i*_, *E*_*i*_, *I*_*i*_, *H* and *R*, with *i* = 1, 2, the fractions of individuals susceptible, exposed, infected, hospitalized and recovered, respectively. Subgroup 1 does not practice social distancing, while Subgroup 2 adopts some level of distancing. From now on, we will refer to these groups as isolated or not, with social distancing or without social distancing, and adopting or not Non-Pharmacological Mitigation Strategies for Diseases (NPMSD).

This approach allows for a detailed analysis of the impact of NPMSD, mainly in relation to social distancing measures. With the inclusion of two levels of social behaviour, the models are able to capture behavioural heterogeneity, addressing more realistic scenarios [11]. In all models, these strategies are represented by dividing the compartments into isolated and non-isolated groups, allowing the evaluation of the impact of different levels of adherence to prevention measures on the dynamics of the disease [3].

### Model 1

In this model, we will assume a hypothetical situation in which the total population (*N*) is subdivided into classes of Susceptible (*S*), Exposed (*E*), Infected (*I*) and Recovered (*R*) individuals. Furthermore, each of the classes of susceptible, exposed, and infected will be subdivided into two subgroups: a group formed by those individuals who practice distancing measures voluntarily or involuntarily (impositions due to health issues), while the second group includes individuals who, voluntarily or involuntarily (social and economical reasons and relationships), do not practice social distancing during a disease outbreak.

The total population is *N* = *S*_1_ + *S*_2_ + *E*_1_ + *E*_2_ + *I*_1_ + *I*_2_ + *R*, where *S*_1_ represents the Susceptible individuals that are not isolated; *E*_1_: non-isolated Exposed; *I*_1_: non-isolated Infected; *S*_2_: isolated Susceptible individuals; *E*_2_: isolated Exposed; *I*_2_: isolated Infected; and *R*: Recovered individuals. In this case, there is no vital dynamic, that is, the population growth rate is considered to be zero, a characteristic usually adopted for the modelling of a shorter epidemic in time when we assume that birth-and-death rates are equal, resulting in a zero population growth.

The resulting model is a version of the SEIR model with the population divided into two levels of behaviours that interact with each other during the epidemic. Using the classic SEIR model as a basis and considering the above hypotheses, the dynamics of disease transmission that we proposed is represented using the flow chart presented in Fig 1.

**Fig 1.**
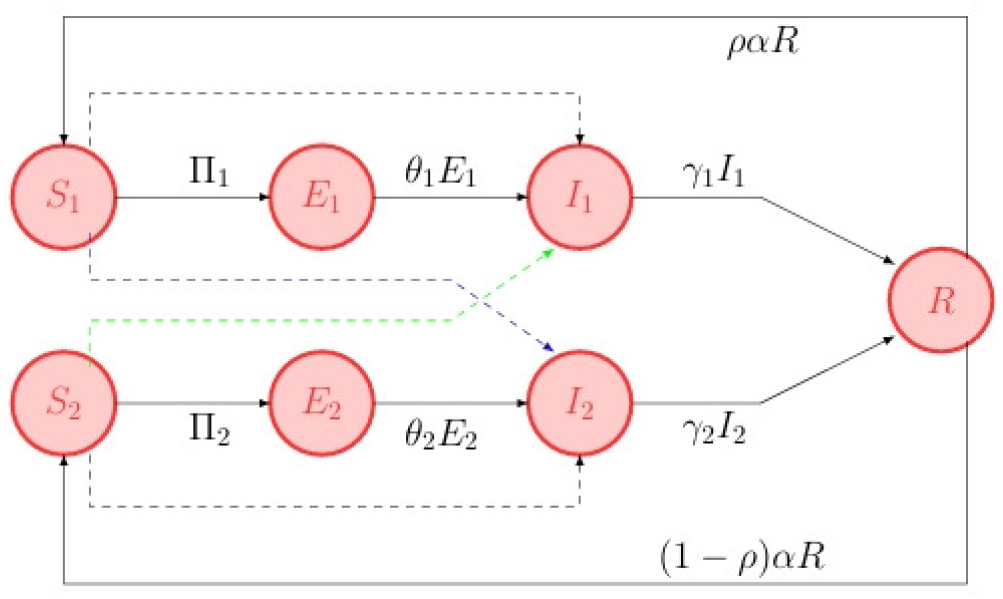
Flow diagram of Model 1, illustrating the interaction between Groups 1 and 2 as well as their internal dynamics. The solid arrows represent the dynamics between the compartments, while the dashed arrows indicate the contacts between the compartments of the same group and of different groups.

Considering Π_1_ and Π_2_ as the expressions that represent all contacts between susceptible and infected in the two groups considered, we have

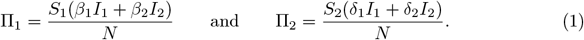

In this model, let us assume that, in the susceptible classes, there are individuals who can contract the disease and, in the class of exposed persons, there are individuals who have contracted the disease but are in the latency period, that is, they do not yet transmit the disease. Classes of infected people transmit the disease when they come into contact with classes of susceptible ones, and in the recovered compartment are individuals with temporary immunity.

Contact between non-isolated susceptible individuals (*S*_1_) and non-isolated infected individuals (*I*_1_) occurs at a rate *β*_1_, and between non-isolated susceptible individuals (*S*_1_) and isolated infected individuals (*I*_2_), at a rate *β*_2_. For susceptible isolates (*S*_2_), contact with isolated infected individuals (*I*_2_) occurs at a rate *δ*_2_, and with non-isolated infected individuals (*I*_1_), at a rate *δ*_1_. The parameters *β*_*i*_ and *δ*_*i*_, with *i* = 1, 2 represent the transmission efficiency of the virus between the susceptible groups and the mentioned infected persons. The values of *β*_*i*_ and *δ*_*i*_ vary between 0 and 1, as the extreme cases of *β*_*i*_ = *δ*_*i*_ = 0 or *β*_*i*_ = *δ*_*i*_ = 1 would indicate both the total absence or the certainty of transmission, respectively, both of which are very unlikely scenarios.

We also consider that infected persons, both non-isolated and isolated, can recover at rates *γ*_1_ and *γ*_2_, respectively, and evolve into the recovered (or removed) compartments. Recovered individuals have temporary immunity and, after a time 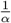, return to the susceptible compartment at the rate *α*. The probability of an individual, after the immunity period, progressing to the compartment *S*_1_, that is, without isolation measures, is *ρ*, while (1 *− ρ*) is the probability of the individual going to the compartment *S*_2_, which adopts measures to mitigate the disease and which we identify as “isolated” as previously indicated.

Therefore, we use the following parameters to describe the dynamics proposed in this model: *β*_1_ and *β*_2_: transmission rate per contact of *S*_1_ with classes *I*_1_ and *I*_2_; *δ*_1_ and *δ*_2_: transmission rate per contact of *S*_2_ with classes *I*_1_ and *I*_2_; *ρ*: probability of a recovered person, after immunity, becoming *S*_1_; 1 *− ρ*: probability of a recovered person, after immunity, becoming *S*_2_; *θ*_1_ and *θ*_2_: transition rate from exposed *E*_1_ and *E*_2_ to infected *I*_1_ and *I*_2_; *γ*_1_ and *γ*_2_: rate of infected people who recover from the disease; and *α*: rate of loss of temporary immunity.

Therefore, based on the assumptions described previously, the dynamics of the model proposed by Fig 1 can be formulated by the following system of ordinary differential equations:

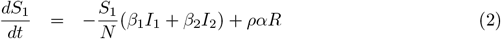

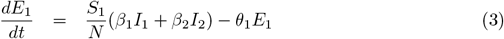

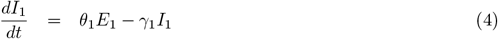

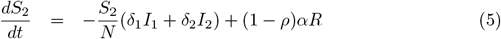

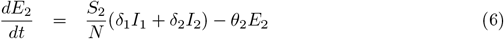

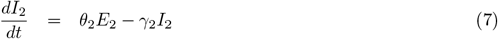

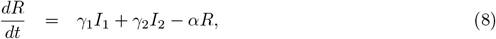

where the initial conditions are: 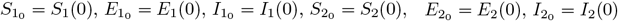 and *R*_0_ = *R*(0).

#### Numerical Simulations of Model 1

Fig 2 shows the dynamics of susceptible populations (*S*_1_ and *S*_2_), exposed populations (*E*_1_ and *E*_2_), infected (*I*_1_ and *I*_2_) and recovered (*R*) populations over 300 days. The population *S*_1_ (without social distancing) decreases rapidly due to increased exposure to the virus, while *S*_2_ (with distancing) shows a slower reduction, reflecting the effectiveness of this measure. Those exposed and infected show initial peaks followed by stabilization, and those recovered grow steadily, suggesting a possible endemic equilibrium. The curves reinforce the importance of social distancing in controlling the spread of the disease.

**Fig 2.**
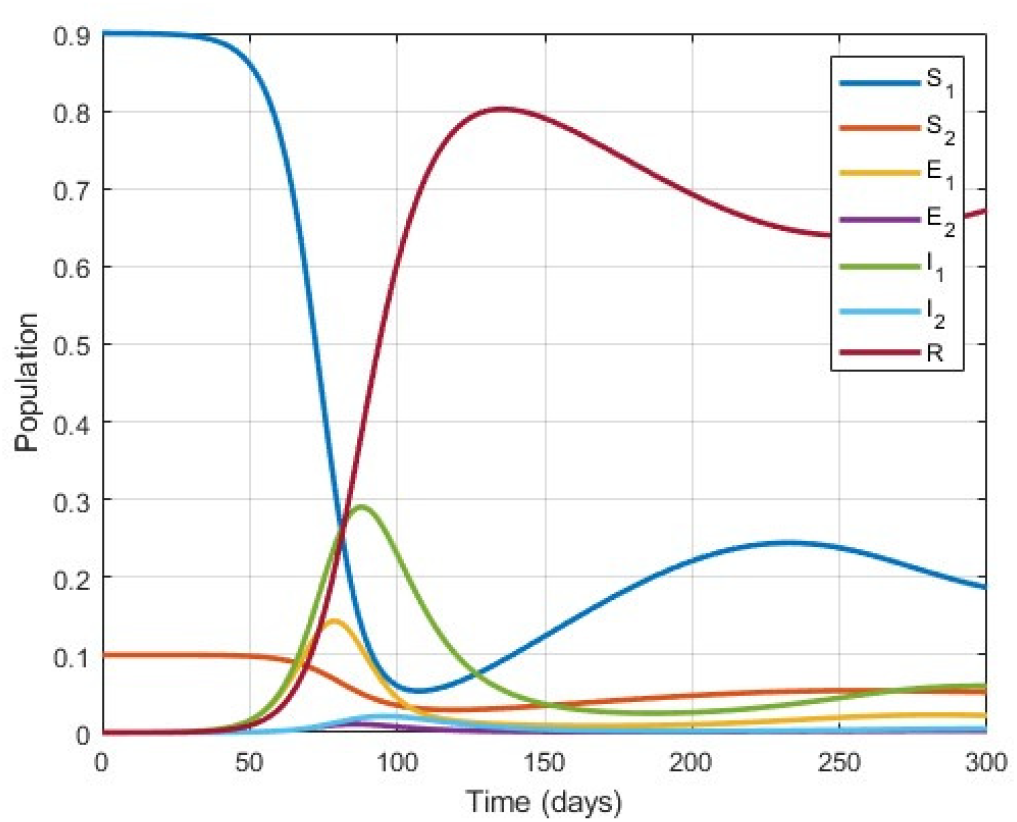
Dynamics of the proposed model. We used the parameters described before and initial conditions *S*_1_ = 0.9, *S*_2_ = 0.1, and *ρ* = 0.9.

Fig 3 shows the aggregate dynamics of susceptible (*S*), exposed (*E*), infected (*I*) and recovered (*R*) over 300 days. The susceptible curve (*S* = *S*_1_ + *S*_2_) decreases rapidly due to the spread of the disease and the effectiveness of the social distancing adopted by *S*_2_. Exposed (*E* = *E*_1_ + *E*_2_) and infected (*I* = *I*_1_ + *I*_2_) individuals show initial peaks, with the highest number of active cases in the first 50 days, when more than 20% of the population were infected, followed by stabilization. The recovered curve (*R*) grows steadily, indicating the recovery of individuals over time.

**Fig 3.**
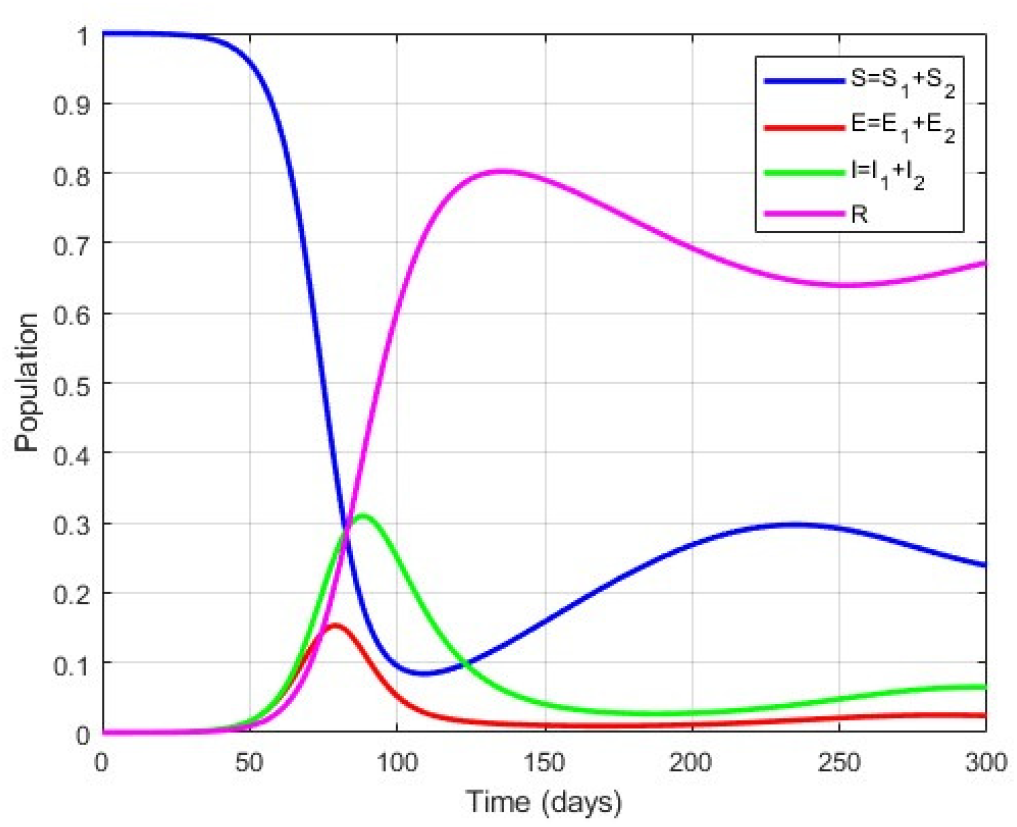
Aggregate dynamics of susceptible populations (*S*), exposed (*E*), infected (*I*) and recovered (*R*) populations over 300 days. We used the parameters described before and initial conditions *S*_1_ = 0.9, *S*_2_ = 0.1, and *ρ* = 0.9.

Fig 4 shows the evolution of active cases of infected 1 (*I*_1_) and infected 2 (*I*_2_), represented by the red and blue curves, respectively. *I*_1_ shows a strong initial peak followed by a smaller wave, while *I*_2_ shows a single later and more intense peak. With initial conditions of *S*_1_(0) = 0.9 and *S*_2_(0) = 0.1, the higher proportion of susceptible in *S*_1_ drives the early peak in *I*_1_, while the lower proportion in *S*_2_ contributes to the later peak in *I*_2_. These patterns reflect the influence of population heterogeneity and highlight the importance of tailored public health measures to control transmission.

**Fig 4.**
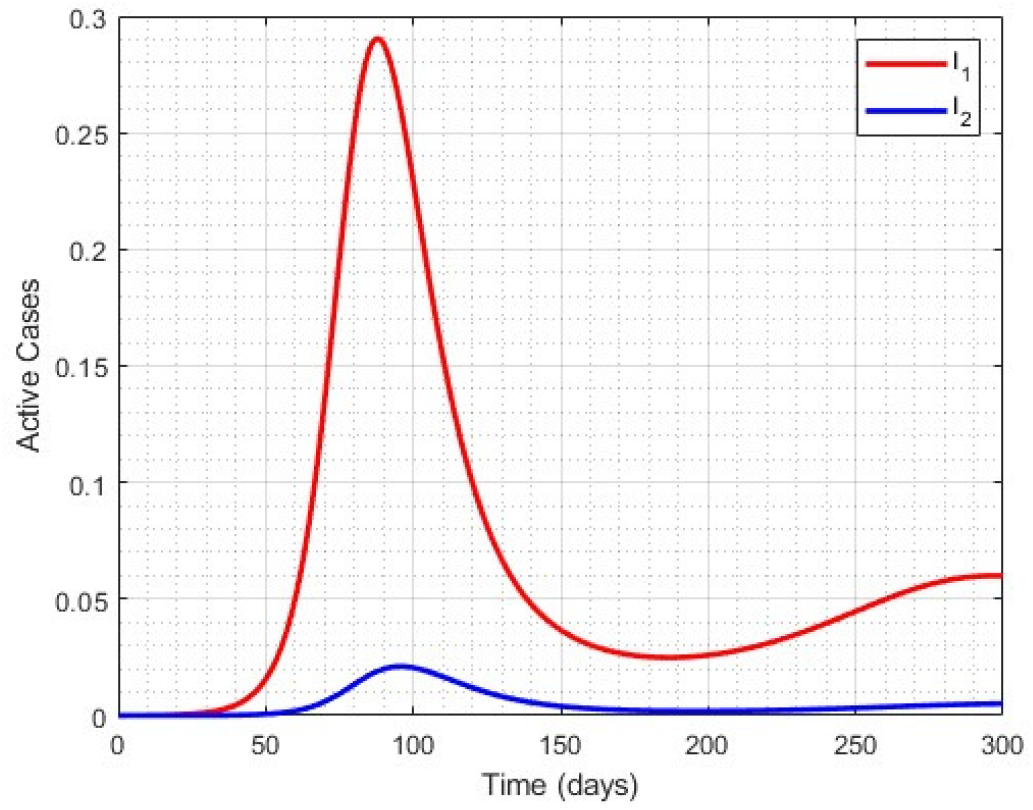
Comparison of the evolution of active cases (*I*_1_) and (*I*_2_). We used the initial conditions *S*_1_ = 0.9 and *S*_2_ = 0.1, and parameters *θ*_1_ = *θ*_2_ = 2.174 *×* 10^*−*1^day^*−*1^, *γ*_1_ = 1.25 *×* 10^*−*1^day^*−*1^, *γ*_2_ = 1.538 *×* 10^*−*1^day^*−*1^ and *α* = 6.667 *×* 10^*−*3^day^*−*1^.

Fig 5 shows four epidemic simulation scenarios, highlighting the impact of non-pharmacological strategies, such as social distancing, on the dynamics of the disease. Each graph illustrates a population divided into proportions of individuals who do not follow (*S*_1_) and who follow (*S*_2_) these strategies. In graph (a), with *S*_1_ = 0.9 and *S*_2_ = 0.1, there is a rapid drop in susceptible individuals and sharp early peaks in exposed and infected individuals, indicating the rapid spread of the disease among those without precautions.

**Fig 5.**
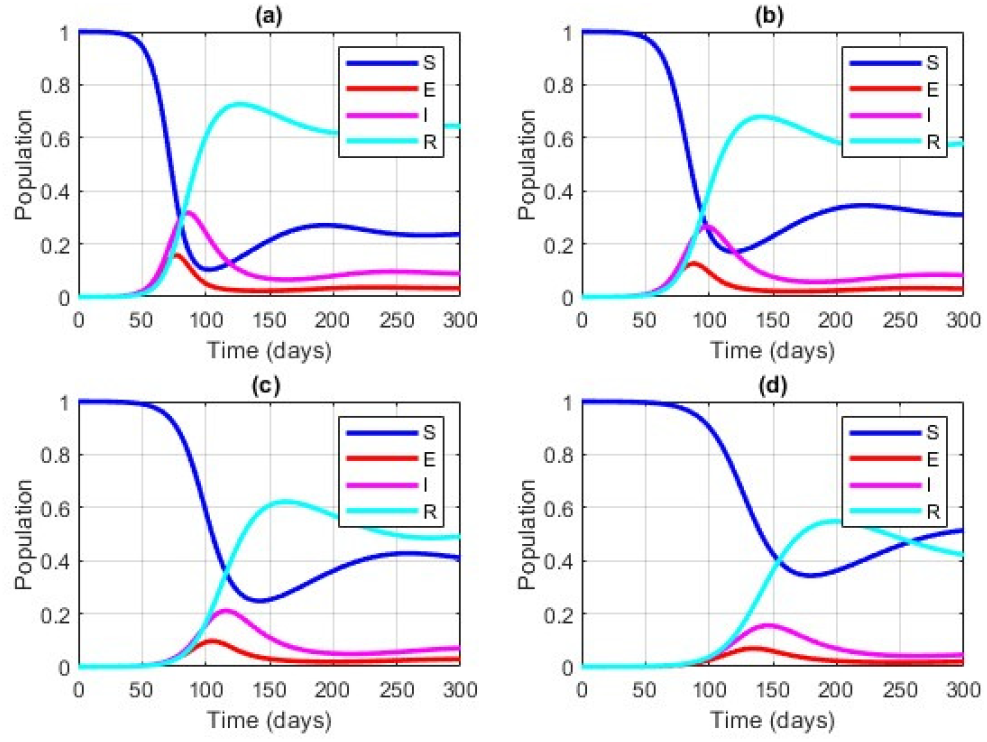
Comparison of model dynamics for initial conditions: (a) *S*_1_ = 0.9 and *S*_2_ = 0.1; (b) *S*_1_ = 0.7 and *S*_2_ = 0.3; (c) *S*_1_ = 0.5 and *S*_2_ = 0.5; (d) *S*_1_ = 0.3 and *S*_2_ = 0.7. We used the parameters *θ*_1_ = *θ*_2_ = 2.174 *×* 10^*−*1^day^*−*1^, *γ*_1_ = 1.25 *×* 10^*−*1^day^*−*1^, *γ*_2_ = 1, 538 *×* 10^*−*1^day^*−*1^ and *α* = 6, 667 *×* 10^*−*3^day^*−*1^.

In graphs (b), (c) and (d), the progressive increase in the proportion of *S*_2_ (*S*_2_ = 0.3, 0.5, 0.7) and the reduction in *S*_1_ result in slower and lower peaks for the infected, showing more effective control of the spread of the disease. In the case of equal division (*S*_1_ = *S*_2_ = 0.5), the peak is still considerable, but less severe than in scenarios with less adherence to mitigation strategies.

In the scenario in (d), with the highest adherence to social distancing (*S*_2_ = 0.7), the peak of infected people is significantly delayed and flattened. This result reflects the effectiveness of social distancing in slowing down transmission and minimizing the impact of an epidemic, confirming the importance of these measures in controlling the spread of a virus.

In summary, Fig 5 illustrates how an increase in adherence to non-pharmacological strategies, such as social distancing, can effectively flatten the epidemiological curve, reducing the number of simultaneously infected people and distributing the demand for health resources over time, which is key to avoiding overloading health systems.

Fig 6 shows the evolution of the percentage of infected (*I*) over time in different initial scenarios, reflecting the impact of compliance with non-pharmacological strategies such as social distancing. Scenarios with a higher initial proportion of *S*_1_ (lower adherence to distancing) result in higher and earlier peaks of infection, indicating accelerated transmission due to the greater number of contacts between susceptible individuals. On the other hand, scenarios with a higher proportion of *S*_2_ (greater adherence to the strategies) show later and flatter peaks, showing a reduction in the transmission rate.

**Fig 6.**
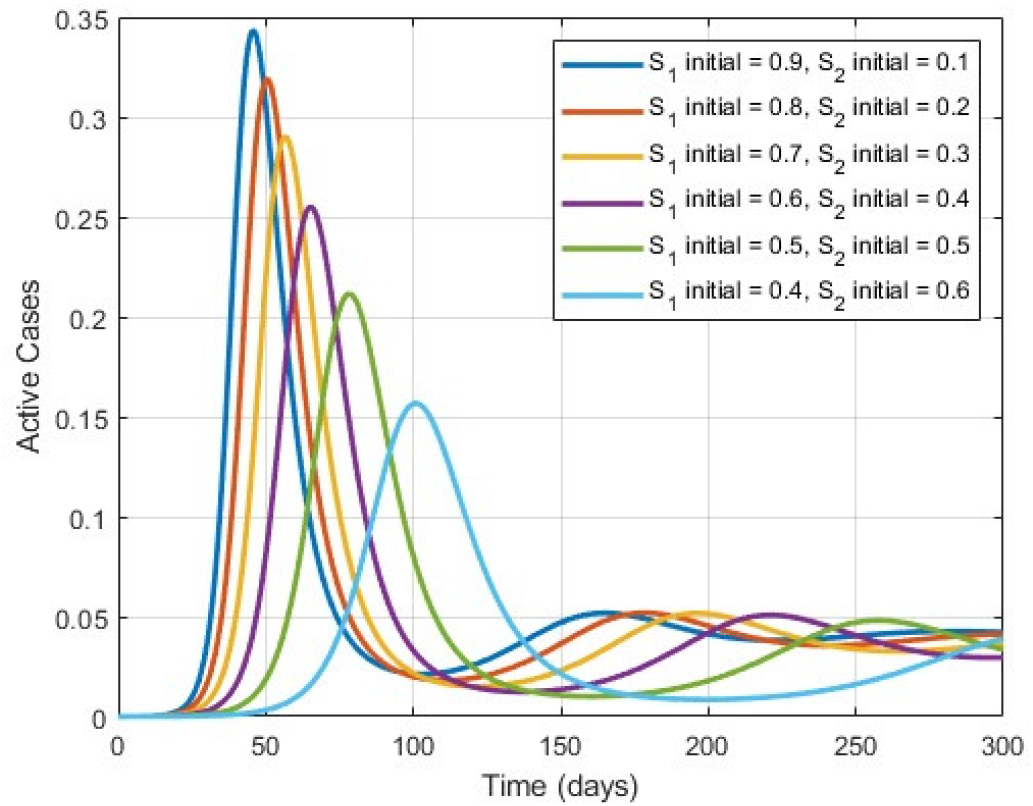
Comparison of active cases of the disease for different initial conditions. We used the parameters *θ*_1_ = *θ*_2_ = 1.9231 *×* 10^*−*1^day^*−*1^, *γ*_1_ = 7.143 *×* 10^*−*1^day^*−*1^, *γ*_2_ = 8.143 *×* 10^*−*1^day^*−*1^ and *α* = 5.667 *×* 10^*−*3^day^*−*1^.

These results highlight the effectiveness of mitigation strategies in containing epidemics, showing that the preventive behaviour adopted by the population can slow and limit the spread of infectious diseases. The figure reinforces the importance of public policies that promote social distancing as a crucial tool for controlling outbreaks and minimizing the impact of epidemics.

Fig 7 shows the evolution of the accumulated infected cases (*I*_1_ + *I*_2_) for different initial combinations of *S*_1_ and *S*_2_. Scenarios with a higher initial proportion of *S*_1_ (e.g. *S*_1_ = 0.9, *S*_2_ = 0.1) show a faster growth in cases at the start of the epidemic, due to the higher transmission rate associated with the population that does not adopt social distancing measures. In contrast, as the proportion of *S*_2_ increases, the growth in cumulative cases becomes slower, reflecting the positive impact of non-pharmacological measures in reducing the spread of the disease.

**Fig 7.**
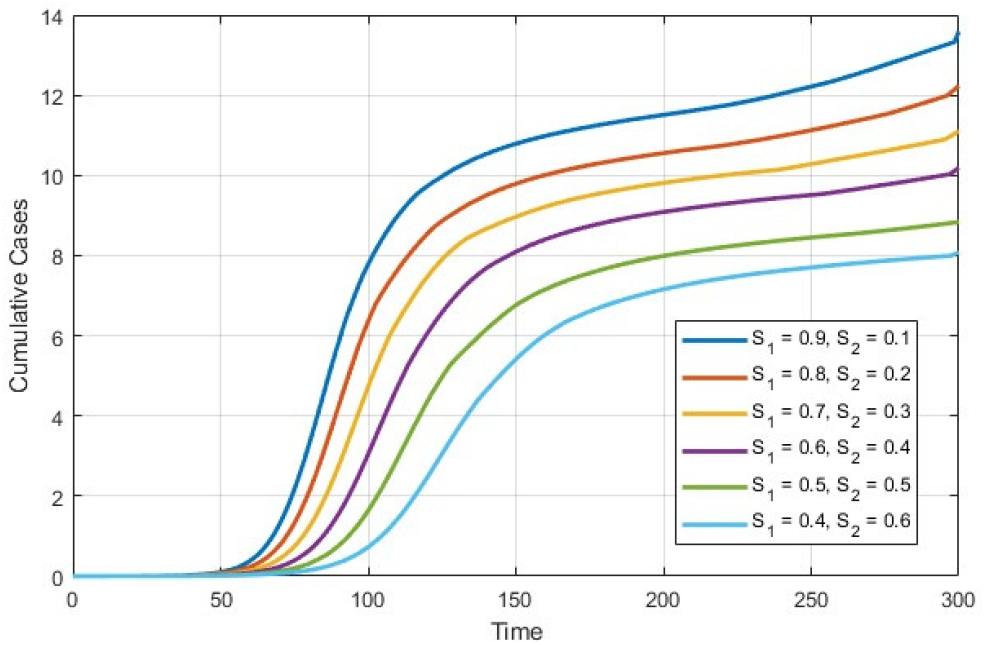
Evolution of cumulative infected cases (*I*_1_ + *I*_2_) with initial conditions of *S*_1_ and *S*_2_. We used the parameters *θ*_1_ = *θ*_2_ = 2.174 *×* 10^*−*1^day^*−*1^, *γ*_1_ = 1.25 *×* 10^*−*1^day^*−*1^, *γ*_2_ = 1.538 *×* 10^*−*1^day^*−*1^ and *α* = 6.667 *×* 10^*−*3^day^*−*1^.

The graph highlights how initial conditions influence the dynamics of the epidemic. The distribution between *S*_1_ and *S*_2_ directly affects the speed of transmission and the total number of cases accumulated. These results reinforce the importance of intervention strategies that encourage increased adherence to preventive measures, showing how changing population behaviour can slow the spread and reduce the impact of the epidemic.

#### Model 2

In this model, we will expand Model 1 to incorporate vital dynamics, taking into account births and deaths in the population, a necessity if one is to consider reasonably longer timespans in the modelling effort. In addition, we will introduce a Hospitalized (*H*) compartment, reflecting the severity of the illness that leads to the need for hospitalization.

This model considers exponential population growth, following the Malthus law, in which the growth rate is proportional to the size of the population. It is essential to understand the dynamics of the disease in contexts where population growth affects transmission. Population dynamics directly influences the number of susceptible people and, therefore, the potential for the disease to spread. Recent studies [13] explore the interaction between population dynamics and disease transmission, highlighting the importance of including exponential growth in the modelling of infectious diseases. Furthermore, considering vital dynamics in the model is crucial to capture changes in the population due to births and deaths, which can affect disease transmission and the effectiveness of interventions over time [14].

In addition, including a hospitalized compartment, *H*(*t*), is valid to model the progression of the disease and the demand for health resources, allowing an analysis of the impact of hospitalizations on the dynamics of the disease [15]. Inclusion of this compartment helps to understand the burden on the health system and the need for hospital beds, intensive care, and medical resources. The other variables in Model 3, *S*_1_(*t*), *E*_1_(*t*), *I*_1_(*t*), *S*_2_(*t*), *E*_2_(*t*), *I*_2_(*t*) and *R*(*t*), are the same as those described in Model 1.

As shown in Fig 8, we consider two main direction sequences in the model. The first follows *S*_1_ *→ E*_1_ *→ I*_1_ *→ H → R*, representing the epidemic for those who do not adopt social distancing. The second follows *S*_2_ *→ E*_2_ *→ I*_2_ *→ H → R*, representing the epidemic for those who adopt some level of social distancing. The dynamics of disease transmission are shown in the flow chart in Fig 8.

**Fig 8.**
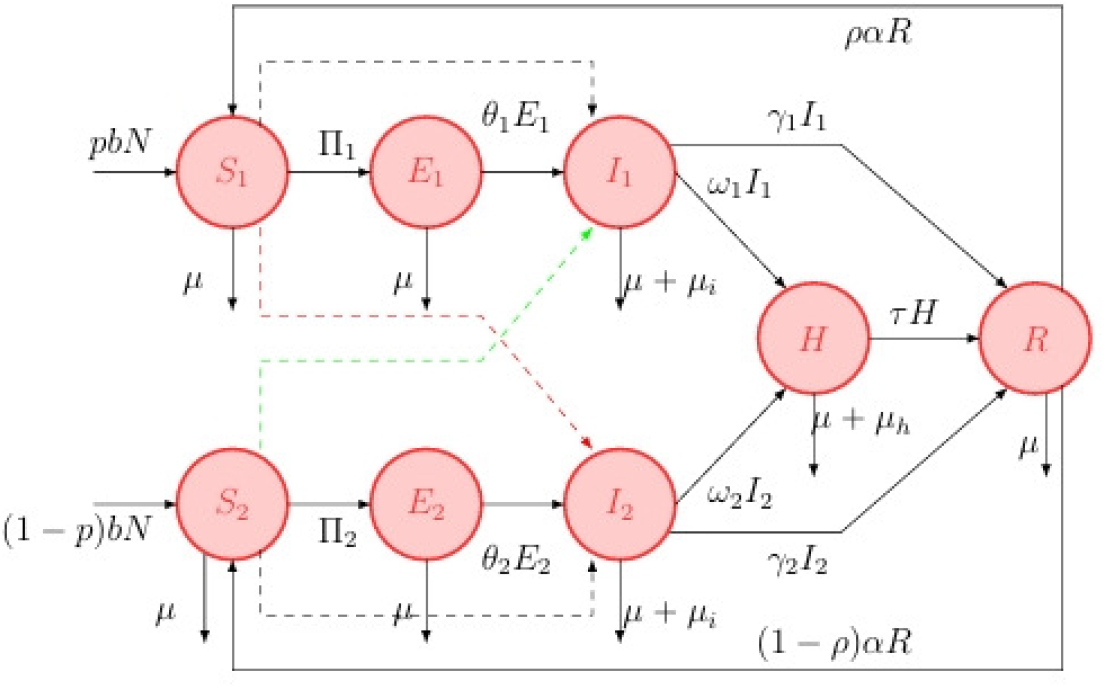
Flow diagram of Model 2, illustrating the interaction between Groups 1 and 2 as well as their internal dynamics.

Taking into account a vital dynamic that includes births and deaths, we also add the birth rate *b*, the natural mortality rate *µ*, the mortality rate due to the disease *µ*_*i*_, and *µ*_*h*_, the mortality rate of those hospitalized due to this same disease. The parameter *p* indicates the fraction of births that enter the compartment of non-isolated susceptible individuals, while (1 *− p*) represents the fraction that enters the compartment of isolated susceptible individuals. The recovered individuals show the same behaviour as described in the previous models. In addition, we have the following parameters: *β*_1_, *β*_2_: contact transmission rate of *S*_1_ with classes *I*_1_ and *I*_2_, respectively; *δ*_1_, *δ*_2_: contact transmission rate of *S*_2_ with classes *I*_1_ and *I*_2_, respectively; *ρ*: probability of the recovered person, after immunity, become *S*_1_; 1 *− ρ*: probability of the recovered person, after immunity, become *S*_2_, *θ*_1_, *θ*_2_: transition rate from exposed *E*_1_ and *E*_2_ to infected *I*_1_ and *I*_2_, respectively; *γ*_1_, *γ*_2_: rate of infected people who recover from the disease; and *α*: rate of loss of temporary immunity.

The model proposed by Fig 8 can then be described by the following system of ordinary differential equations:

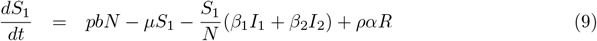

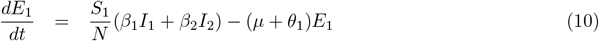

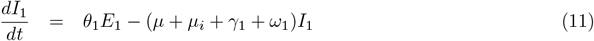

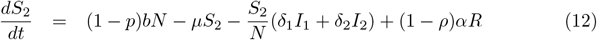

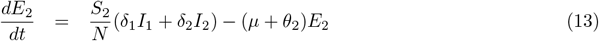

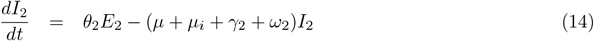

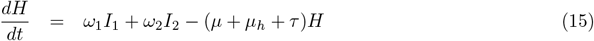

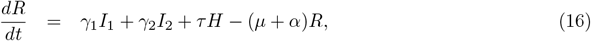

#### Numerical Simulations of Model 2

First, let us analyse the disease-free equilibrium, considering the point *E*_0_ = (*S*_1_, *E*_1_, *I*_1_, *S*_2_, *E*_2_, *I*_2_, *H, R*), with *I*_1_ = 0 and *I*_2_ = 0, and *S*_1_ = 0.8 and *S*_2_ = 0.2. The graph in Fig 9 illustrates the evolution of various population categories over time. At the disease-free equilibrium point, *E*_0_ = (0.8, 0, 0, 0.2, 0, 0, 0), of model 2, which represents a scenario in which the disease is not initially present. The lines representing the susceptible subpopulations, *S*_1_ and *S*_2_, remain constant over time, at 0.8 and 0.2, respectively.

**Fig 9.**
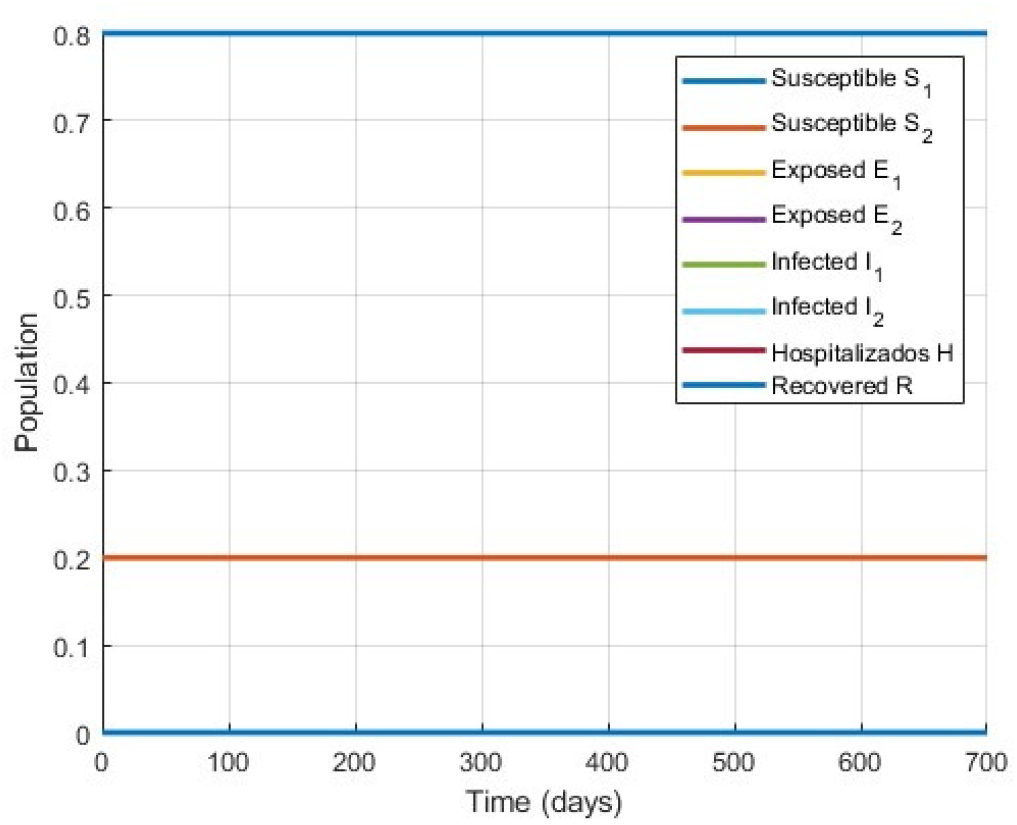
Dynamics of susceptible populations (*S*_1_) and (*S*_2_) at the Disease-free Equilibrium Point.

In summary, the graph shows a disease-free equilibrium state in which the initial population does not experience infection, progression to exposure, infection, or hospitalization. This is consistent with the initial conditions established: a large part of the population is susceptible *S*_1_ = 0.8 and *S*_2_ = 0.2, and there are no individuals initially exposed, infected, hospitalized, or recovered. The absence of any variation in the lines over time confirms that the system remains stable and disease-free throughout the simulation period.

Consider a disease with infection rates given by *β*_1_ = 0.6, *β*_2_ = 0.4, *δ*_1_ = 0.3, *δ*_2_ = 0.15. According to the hypothesis of the model formulation, we have *β*_1_ *> β*_2_ and *δ*_1_ *> δ*_2_, as well as *β*_2_ *> δ*_1_. Suppose that *I*_1_ and *I*_2_ are different from zero, indicating that the disease is present in the population. Using the initial conditions:

*S*_1_(0) = 7.99995 *×* 10^*−*1^, *S*_2_(0) = 1.99995 *×* 10^*−*1^, *I*_1_(0) = *I*_2_(0) = 5.0 *×* 10^*−*6^, and *H*(0) = *R*(0) = 0, and the parameters: *ρ* = 8.0 *×* 10^*−*1^ estimated; *α* = 5.5556 *×* 10^*−*3^ [16]; *p* = 8.0 *×* 10^*−*1^ estimated; *µ* = 3.5 *×* 10^*−*5^ estimated; *µ*_*i*_ = 4.0 *×* 10^*−*4^ estimated; *µ*_*h*_ = 1.0 *×* 10^*−*4^ estimated; *θ*_1_ = *θ*_2_ = 1.9231 *×* 10^*−*1^ [17]; *γ*_1_ = *γ*_2_ = 6.6661 *×* 10^*−*2^ [18]; *ω*_1_ = *ω*_2_ = 1.9 *×* 10^*−*2^ [19]; *τ* = 1.0 *×* 10^*−*1^ [19]; and *b* = 3.55 *×* 10^*−*5^ [20]. The dynamics of the model is illustrated in Fig 10.

**Fig 10.**
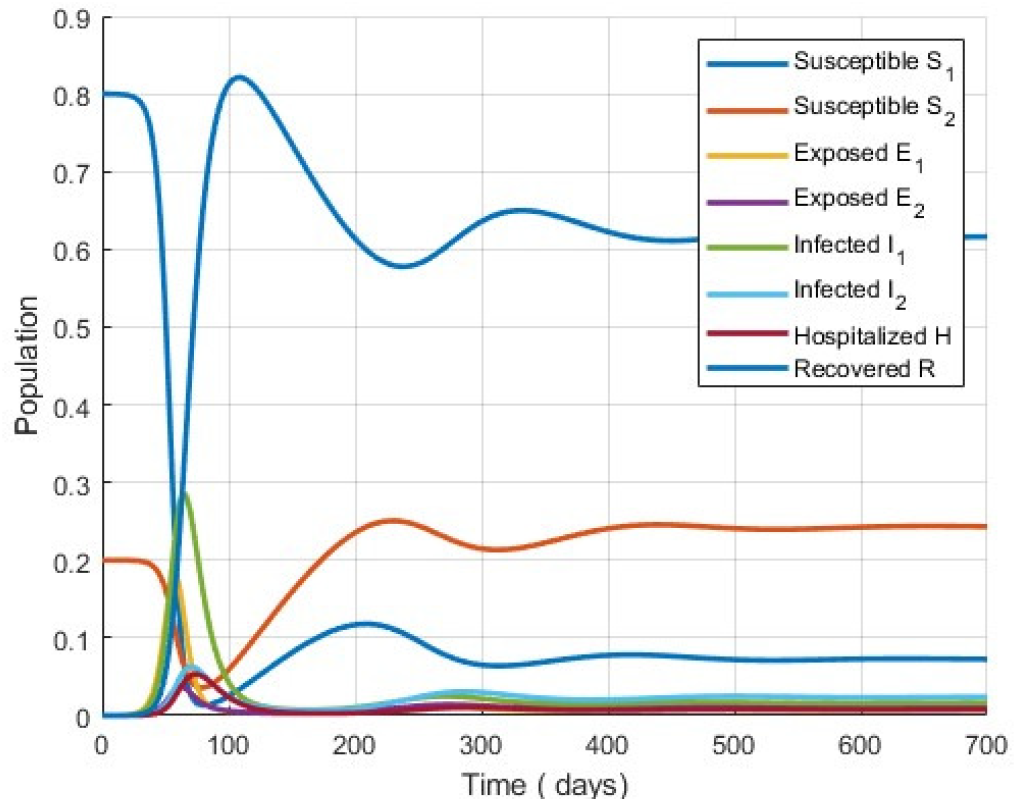
Dynamics of susceptible (*S*), exposed (*E*), infected (*I*) and recovered (*R*) populations over 700 days.

Fig 10 shows the evolution of population categories over time: susceptible (*S*_1_ and *S*_2_), exposed (*E*_1_ and *E*_2_), infected (*I*_1_ and *I*_2_), hospitalized (*H*) and recovered (*R*). The *S*_1_ population decreases rapidly, reaching a peak of infections around day 50 and then stabilizing at a lower level. The population *S*_2_ decreases more slowly, reflecting the impact of containment measures.

For a better understanding of the dynamics of Model 2, Fig 11 shows the temporal evolution of the sums of the different categories over 700 days.

**Fig 11.**
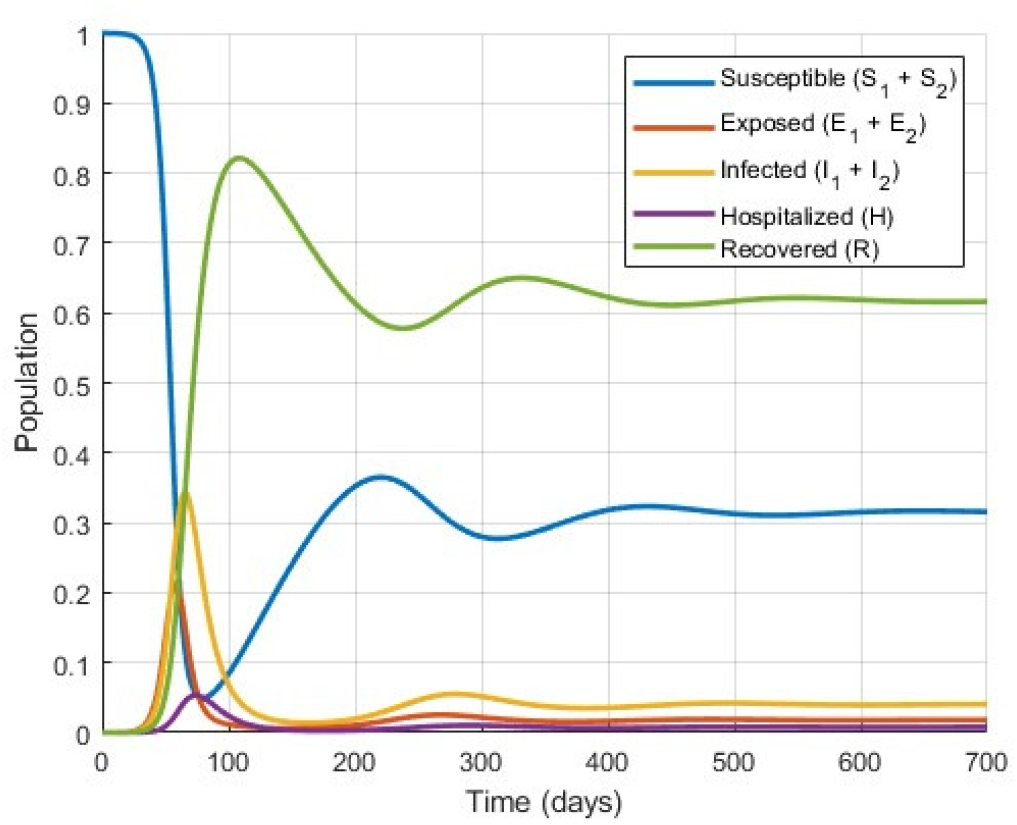
Dynamics of susceptible (*S*), exposed (*E*), infected (*I*) and recovered (*R*) populations over 700 days.

Fig 12 shows how the susceptible population decreases due to infection, while the exposed and infected populations increase. The peak of infection, represented by the sum of those infected (*I*_1_ + *I*_2_), occurs on approximately the 60th day, indicating the time of greatest transmission of the disease.

**Fig 12.**
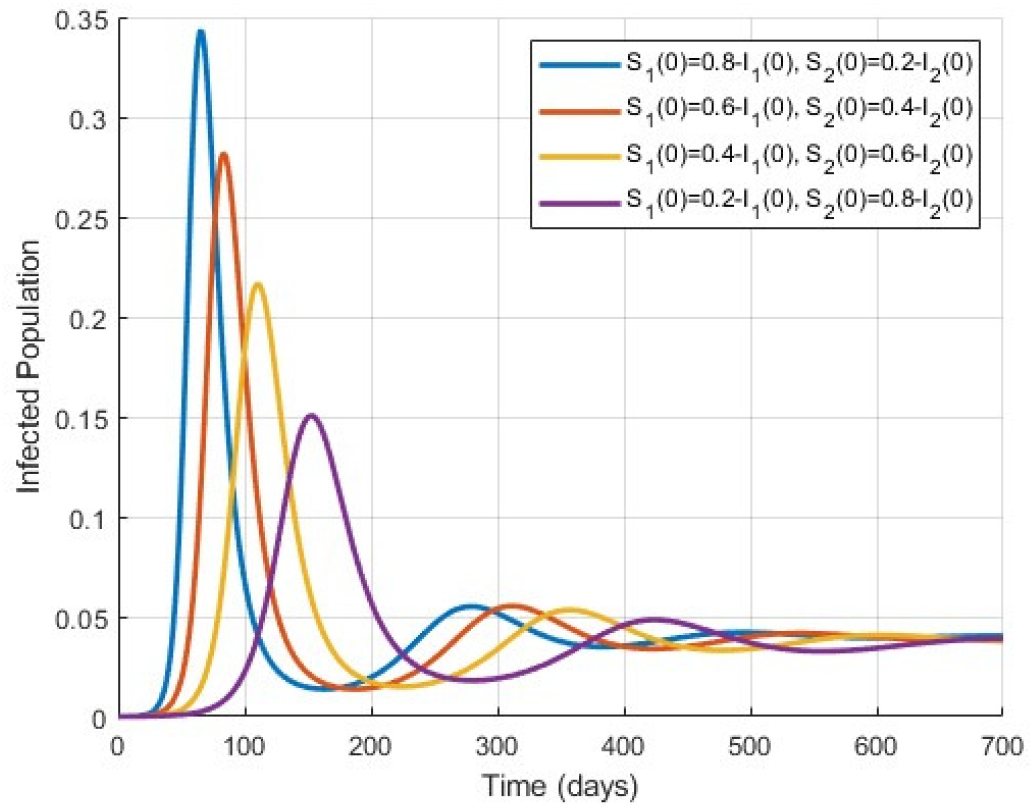
Dynamics of susceptible (*S*), exposed (*E*), infected (*I*) and recovered (*R*) populations over 700 days.

As the outbreak progresses, the hospitalized curve increases, reflecting the growing need for medical care for the severely infected. The population of recovered people grows with time, indicating the recovery rate. Approximately after day *t* = 250, the number of new cases begins to decrease, suggesting that the epidemic is being controlled. At the end of the simulation period, the populations stabilize, reflecting the control of the epidemic and the arrival at a state of equilibrium. The graph suggests that the system has reached stability.

In Fig 13, we analyse the evolution of the hospitalized population (*H*) over 700 days under different initial conditions for susceptible ones (*S*_1_(0) and *S*_2_(0)):

**Fig 13.**
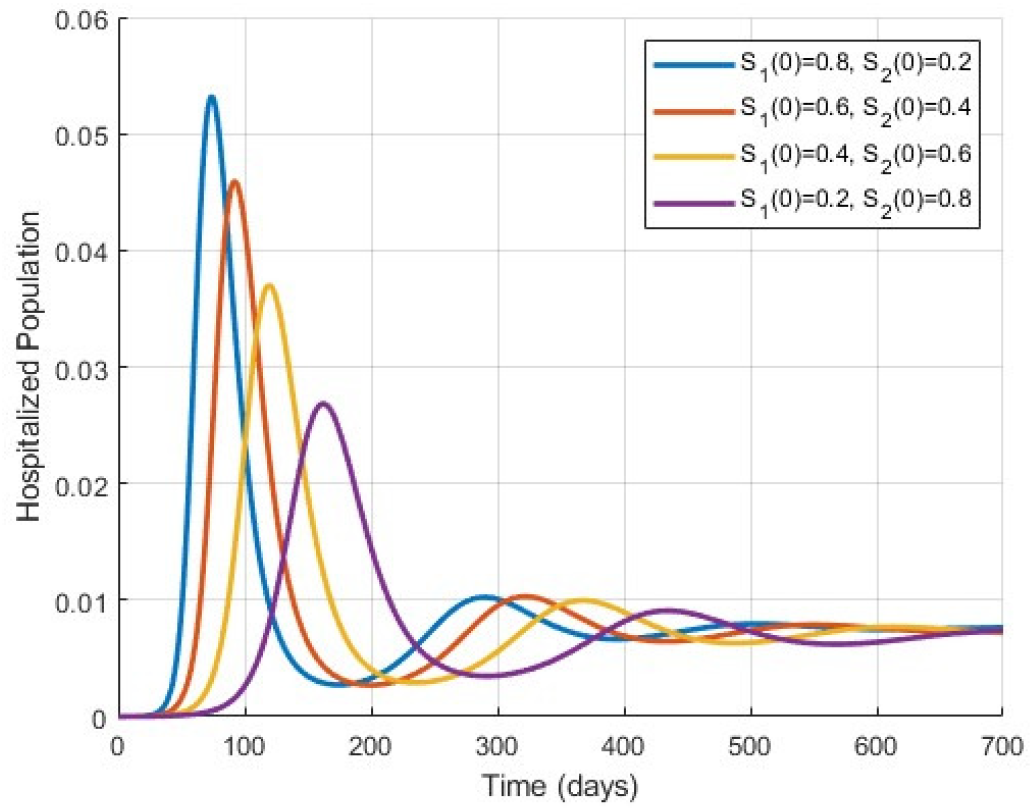
Evolution of the infected population over time with variation of the parameters *α* and *ρ*. Subplots (a) and (b) show the dynamics of infected (*I*_1_ + *I*_2_) for different values of *ρ* : (0.9 *≤ ρ ≤* 0.1). 2.

- *S*_1_(0) = 0.8 and *S*_2_(0) = 0.2: a peak occurs around day 70, followed by a smaller second peak near day 280, and then stabilization;
- *S*_1_(0) = 0.6 and *S*_2_(0) = 0.4: a peak is observed around day 80, with subsequent oscillations before stabilizing;
- *S*_1_(0) = 0.4 and *S*_2_(0) = 0.6: a peak occurs around day 110, followed by a decline and a smaller secondary peak before stabilization;
- *S*_1_(0) = 0.2 and *S*_2_(0) = 0.8: a lower and slightly delayed peak occurs around day 130, with smaller subsequent oscillations.

When comparing these dynamics with Fig 11 (infected population), we observe that the hospitalization peaks follow the infection peaks, which is expected since hospitalizations are a consequence of infections. The transmission rates *β*_1_ = 0.6, *β*_2_ = 0.4, and *δ*_1_ = 0.3, *δ*_2_ = 0.15 directly influence these dynamics.

The observed trends indicate a **trend toward equilibrium** regardless of the initial conditions. However, eigenvalues with small positive real parts (0.0147 *±* 0.0195*i*) suggest a **potential long-term instability**, which could lead to persistent oscillations or the growth of infections within certain subpopulations.

In summary, the graphs illustrate how different containment strategies, represented by the proportions of *S*_1_ and *S*_2_, and modulated by transmission rates, influence disease propagation and hospitalization dynamics over time. This highlights the complexity of the epidemiological model and its stability characteristics.

Fig 14 shows the evolution of the infected population over time, considering two distinct values for the temporary immunity rate (*α*) and various values for the probability of post-immunity behaviour (*ρ*).

**Fig 14.**
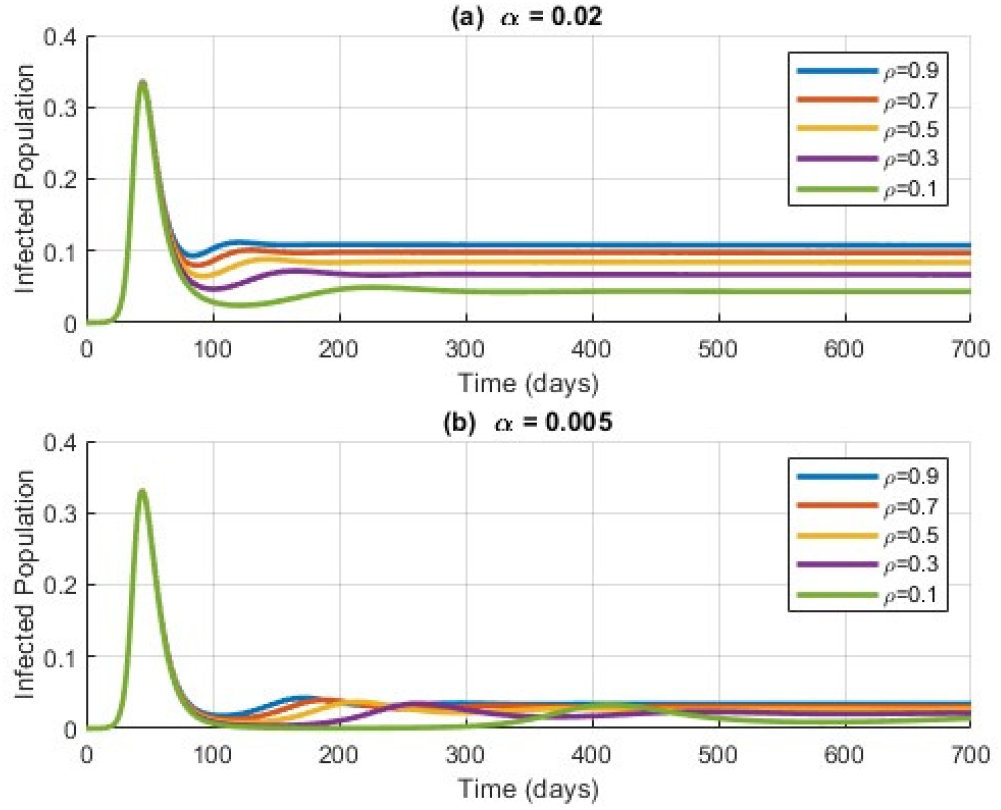
Evolution of the infected population over time with variations in the parameters *α* and *ρ*. Sub-plots (a) and (b) show the dynamics of infected (*I*_1_ + *I*_2_) for different values of *ρ* : (0.9 *≤ ρ ≤* 0.1).

The parameter *α* represents the duration an individual remains immune after recovering from the disease. In contrast, *ρ* denotes the probability that, after the immunity period, an individual returns to the social behaviour of (*S*_1_), while 1 *− ρ* indicates the likelihood of adopting the behaviour of (*S*_2_).

Fig 14 consists of two subplots, each corresponding to a different value of *α*:

1. Sub-plot (a) (*α* = 0.02):
  - A higher temporary immunity rate (*α* = 0.02) results in a shorter immunity period.
  - Higher values of *ρ* lead to faster and higher infection peaks due to the return to non-isolated behaviour.
2. Sub-plot (b) (*α* = 0.005):
  - A lower temporary immunity rate (*α* = 0.005) results in a longer immunity period.
  - Lower values of *ρ* slow the spread of the disease and produce lower infection peaks, reflecting prolonged isolation behaviour.

The probability of post-immunity behaviour (*ρ*) significantly impacts disease dynamics: higher *ρ* values result in faster and higher infection peaks, as individuals return to non-isolated behaviour (*S*_1_), increasing reinfection. In contrast, lower *ρ* values, where individuals adopt isolated behaviour (*S*_2_), help reduce disease spread.

This analysis highlights the importance of incorporating temporary immunity (*α*) and post-immunity behaviour (*ρ*) into models to better understand and develop strategies for controlling infectious diseases.

#### Model 3

In this third model, we will expand Model 2 to include a new compartment for vaccinated individuals (*V*). This addition is extremely important to understand and evaluate the impact of vaccination on disease dynamics and population [21]. Therefore, the total population will be defined by *N* = *S*_1_ + *S*_2_ + *E*_1_ + *E*_2_ + *I*_1_ + *I*_2_ + *H* + *R* + *V*, with *S*_1_, *E*_1_ and *I*_1_ representing the classes of individuals who do not practice social distancing (that is, situations without non-pharmacological strategies) and *S*_2_, *E*_2_ and *I*_2_ representing the classes of individuals who reduce physical proximity (with non-pharmacological strategies).

Inclusion of the vaccine compartment is also essential for evaluating the effect of vaccination on reducing susceptibility to the disease, as well as reducing transmission.

It is important to highlight the interaction between non-pharmacological strategies, such as social distancing, the use of masks, and vaccination strategies. The combination of these and other measures can be important in controlling the spread of the disease and protecting a population, especially in scenarios where vaccination is not universal or the effectiveness of the vaccine is limited [3]. Thus, the model makes it possible to analyse the balance between the different strategies to prevent and control the disease.

In addition, we will introduce into this dynamic a parameter *ϑ* which represents a change in the behaviour of the population during the epidemic period, allowing individuals in group 1 *S*_1_, *E*_1_, *I*_1_ to evolve to group 2 *S*_2_, *E*_2_, *I*_2_ and vice versa at any time during the epidemic, characterizing a change in their social attitude towards the use or not of non-pharmacological strategies to mitigate a certain disease. This change in attitude is quite possible during epidemics, in which the population changes its way of acting according to the time and society’s perception of the risks due to the epidemic. Studies show that attitudes change frequently during epidemics, and people adjust their actions based on risk assessment and the efficiency of protective measures [22, 23].

Based on the assumptions described above, the dynamics of the proposed model presented in Fig 15 can then be described by the following system of ordinary differential equations

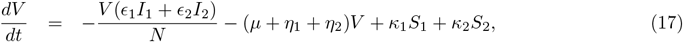

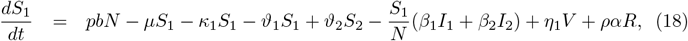

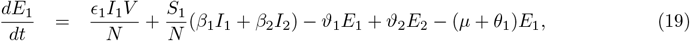

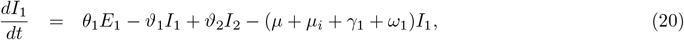

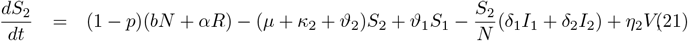

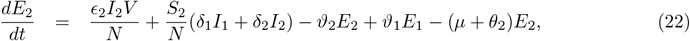

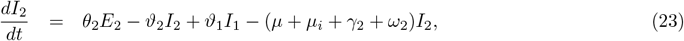

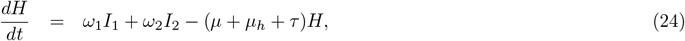

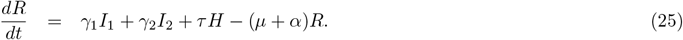

where the population size at time *t* is given by

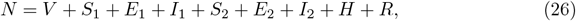

in which we consider 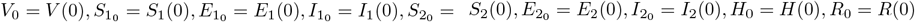 as the initial conditions of this system.

**Fig 15.**
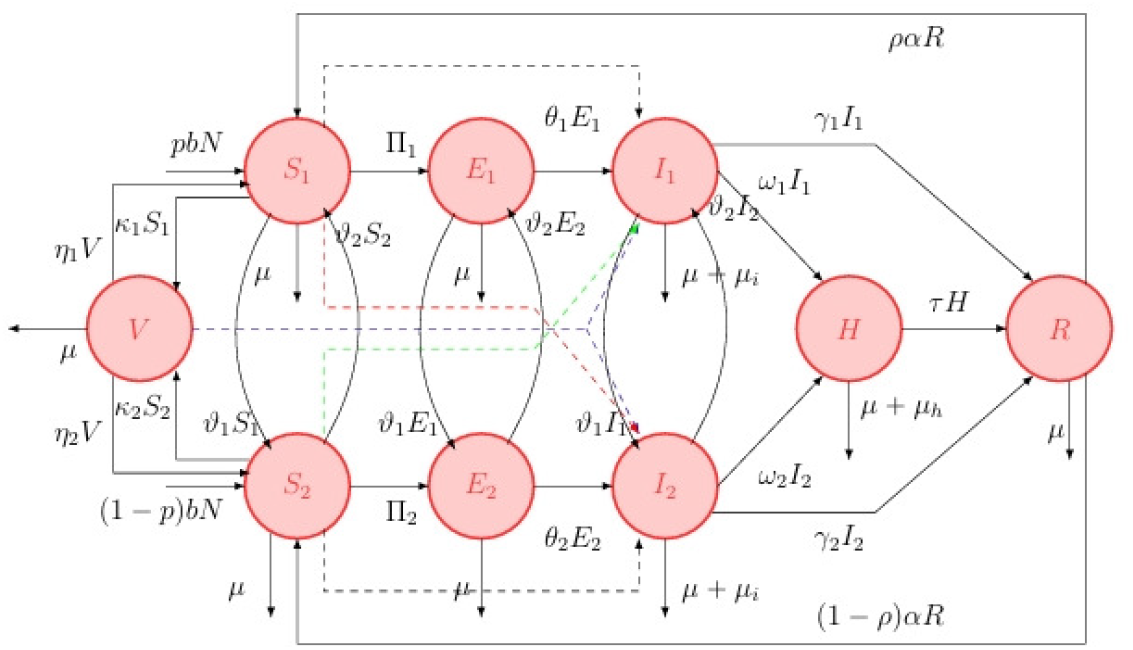
Flow diagram of Model 3, illustrating the interaction between Groups 1 and 2 as well as their internal dynamics.

#### Numerical Simulations of Model 3

We now present simulations of Model 3, which incorporates new aspects of the previous models, as well as the parameter *ϑ*, which represents behavioural changes during the epidemic, and the inclusion of vaccination. The analyses assess the impact of these changes and vaccination on the evolution of the epidemic and the stability of the equilibrium points.

We considered a hypothetical disease with transmission and recovery rates *β*_1_, *β*_2_, *δ*_1_ and *δ*_2_, given the conditions *β*_1_ *> β*_2_, *δ*_1_ *> δ*_2_ and *β*_2_ *> δ*_1_. The parameters used are the same from the simulations of Model 2. Initially, we simulated the absence of vaccination and analysed the behaviour of active cases under three different behaviour change scenarios indicated by *ϑ*.

Fig 16 shows the evolution of active cases over time, considering changes in social behaviour during the outbreak. The simulations analyse three distinct intervals for the changes in behaviour represented by *ϑ*_1_ (greater social distancing) and *ϑ*_2_ (reduced distancing). There is a significant initial peak in active cases due to initial conditions, with a large part of the population in *S*_1_.

**Fig 16.**
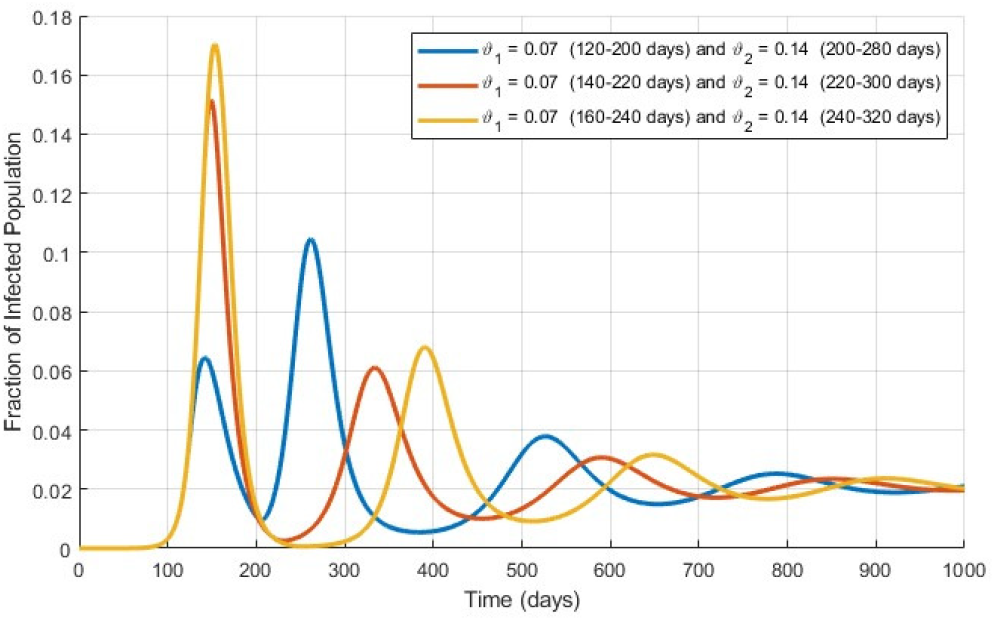
Comparison of the dynamics of active cases (*I*_1_ + *I*_2_) for *ϑ*_1_ = 0.07 in the intervals (120 *< t <* 200), (140 *< t <* 220), (160 *< t <* 240) and *ϑ*_2_ = 0.14 in the intervals (200 *< t <* 280), (220 *< t <* 300), (240 *< t <* 320).

Changes in greater social distancing (*ϑ*_1_) occur between the 120th and 200th days, the 140th and 220th days, and the 160th and 240th days, while reversals to less distancing (*ϑ*_2_) occur between the 200th and 280th days, 220th and 300th days, and the 240th and 320th days. These dynamics reflect the population’s behavioural responses to the outbreak, with isolation measures being adopted and relaxed at specific times.

The introduction of *ϑ*_1_ temporarily reduces active cases, as evidenced by the drop in the curves after the initial peaks. On the other hand, *ϑ*_2_ generates additional outbreaks, reflecting a lower adherence to social separation. The oscillations in the curves indicate secondary outbreaks due to the alternation between social behaviours and possible reinfections.

The graph thus highlights the relevance of the parameters *ϑ*_1_ and *ϑ*_2_ in analysing the dynamics of directly transmitted diseases. Social distancing measures (*ϑ*_1_) help temporarily contain cases, while relaxing these measures (*ϑ*_2_) causes new increases in cases of infection.

Fig 17 is a more detailed version of the previous figure, illustrating the evolution of active cases of infection over time (in days), with different conditions for changes in social behaviour represented by *ϑ*_1_ and *ϑ*_2_. These graphs allow a clear analysis of infection patterns in the face of changes in social behaviour during the epidemic outbreak. In all cases, the probability *ρ* that an individual returns to a behaviour without social separation (*S*_1_) or adopting mitigation measures (*S*_2_) is the same.

**Fig 17.**
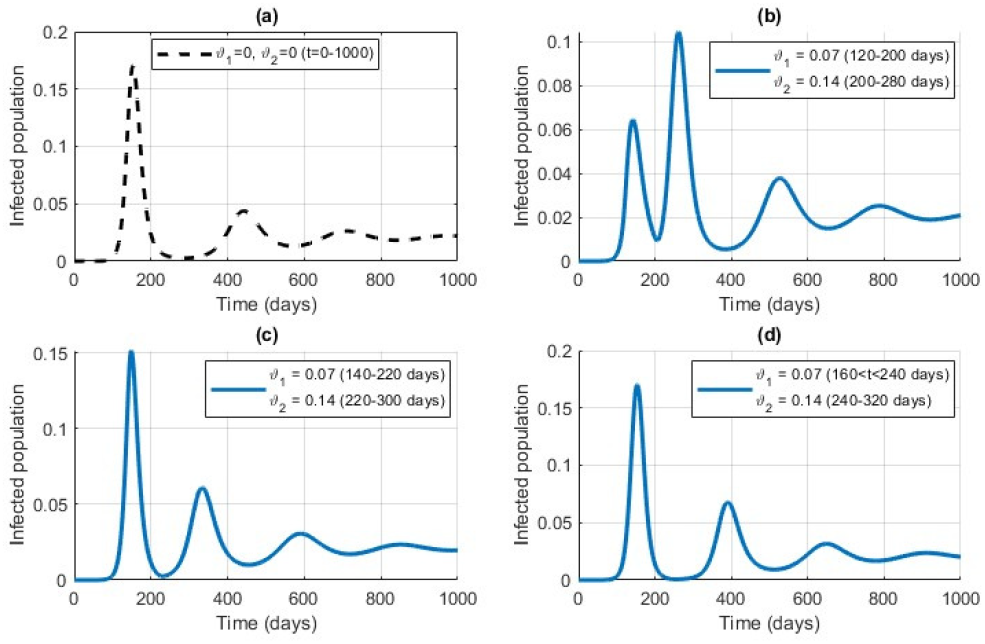
Detailed dynamics of infected with *ϑ*_1_ = *ϑ*_2_ = 0 for all *t* and *ϑ*_1_ = 0.07 in the intervals (120 *< t <* 200), (140 *< t <* 220) and (160 *< t <* 240) and *ϑ*_2_ = 0.14 in the intervals (200 *< t <* 280), (220 *< t <* 300) and (240 *< t <* 320).

Fig 17 shows the evolution of infection cases in different scenarios of changes in social behaviour, detailing the phases of introduction (*ϑ*_1_) and reversal (*ϑ*_2_) of social distancing measures. In graph (a), we present the situation without change in social behaviour (*ϑ*_1_ = 0 and *ϑ*_2_ = 0), there is a significant initial peak followed by oscillations and secondary surges. In graph (b), we have *ϑ*_1_ = 0.07 implemented between 120 and 200 days and *ϑ*_2_ = 0.14 between 200 and 280 days, there is a temporary reduction in cases, followed by new outbreaks after the reversal. In graph (c), the changes made in subsequent periods (*ϑ*_1_ = 0.07 between 140 and 220 days and *ϑ*_2_ = 0.14 between 220 and 300 days) show a similar impact, with oscillations influenced by social distancing. Finally, in graph (d), we present a situation similar to graph (c), but with the changes applied later, strengthening the role of *timing* in controlling the epidemic.

The analysis highlights that social distancing measures (*ϑ*_1_) temporarily reduce active cases, while their reversal (*ϑ*_2_) leads to new outbreaks. By maintaining the same probability of migration (*ρ*) between social behaviours, the graphs highlight the importance of well-planned and timed strategies for effective control of the epidemic.

Now let us analyse the model considering the existence of a vaccine against this hypothetical disease and evaluate the scenario with vaccination started at different times. Looking at the graph in Fig 18, we have the evolution of active cases of infected people in the model presented, for different conditions of vaccination initiation: no vaccination, vaccination initiated in the fourth month, in the sixth month, and in the 12th month.

**Fig 18.**
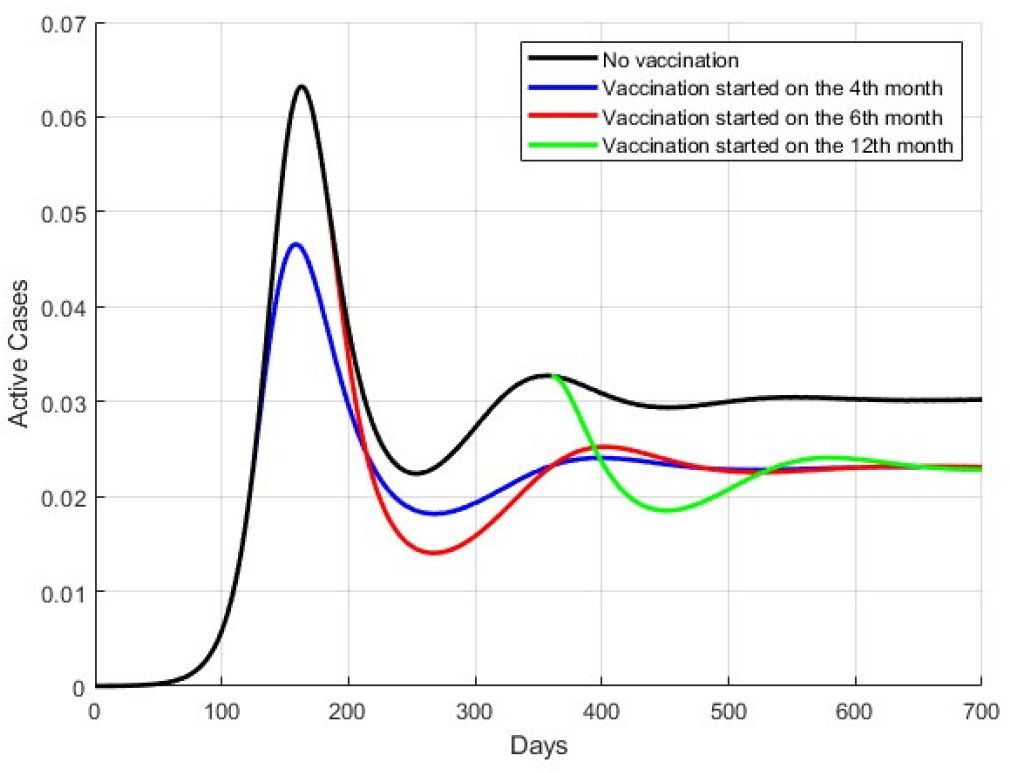
Comparative dynamics of the evolution of active cases without vaccination and with vaccination initiated in the 4th, 6th and 12th months.

Without vaccination, there is an initial peak around the 160th day, followed by a decrease and smaller oscillations. This scenario shows the natural evolution of the epidemic without intervention, with instability and continuous cycles of infection.

With vaccination starting in the fourth month, the peak in infections occurs around the 140th day, is lower, and stability is reached more quickly. In the case of vaccination starting at month 6, the peak occurs around day 160, coinciding with the peak without vaccination, but still significantly reducing active cases. When vaccination begins in the 12th month, the peak also occurs around the 160th day, but is high and stability is achieved later.

By comparing the scenarios, it can be seen that vaccination starting around the peak without vaccination has less effect on reducing active cases. However, vaccination, in any of the suggested scenarios, still produces a decrease in the number of cases and a stabilization of the model at lower levels of active cases than without vaccination. Vaccination started earlier, before the expected peak, has a more significant impact on reducing the peak of infections and stabilizing cases over time. This highlights the importance of earlier interventions to effectively control the spread of infection and avoid high peaks in active cases.

In Fig 19, we observe the evolution of the model’s active infected cases (15) in detail for different vaccination start conditions: no vaccination and vaccination started at the fourth, sixth, and 12th months.

**Fig 19.**
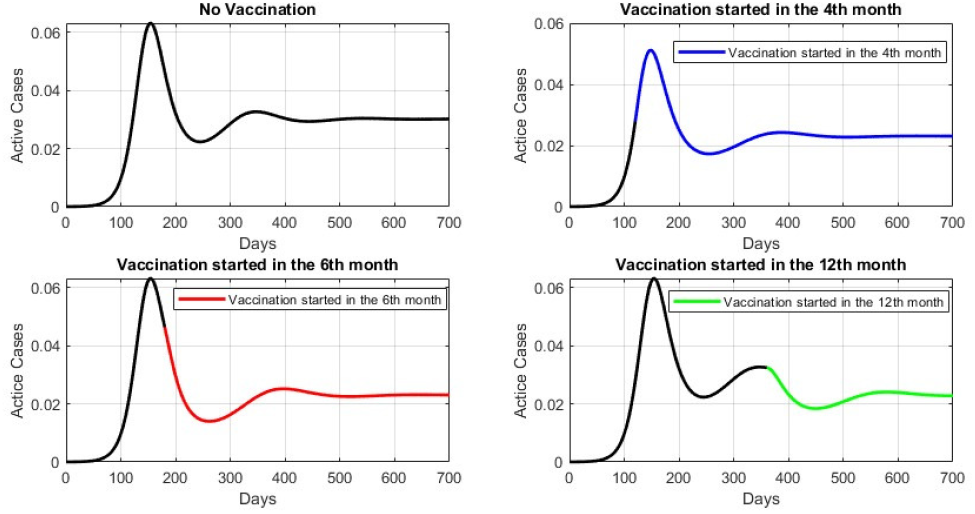
Detailed comparative dynamics of the evolution of active cases without vaccination and with vaccination initiated in the 4th, 6th and 12th months.

Vaccination in the fourth month rapidly reduces active cases, stabilizing the system earlier. Starting vaccination in the sixth month, the reduction in cases is less pronounced and stabilization occurs later. Vaccination in the 12th month results in a reduction in cases, but with less impact and delayed stability.

In all conditions, the probability of returning to the compartment *S*_1_ is *ρ* = 0.8 and for *S*_2_ it is 1 *− ρ*. The early introduction of vaccination promotes more efficient control and rapid stabilization of the epidemic.

The graph in Fig 20 shows the number of cumulative infected cases over time for four different conditions: no vaccination, vaccination started in the fourth month, vaccination started in the sixth month, and vaccination started in the 12th month.

**Fig 20.**
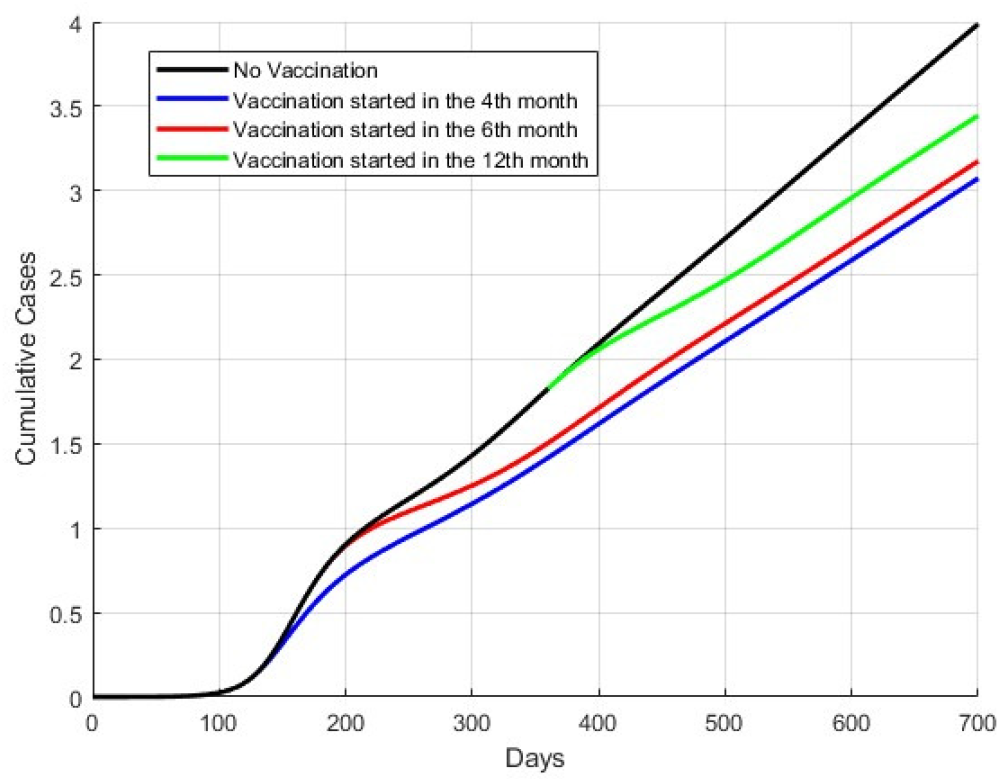
Evolution of the accumulated cases for the different conditions of vaccination onset.

Analysing the graph presented in Fig 20, we can observe that when there is no vaccination, the curve shows a constant and rapid growth in the number of accumulated cases, reaching a significantly high number during the simulation time. This shows the continuous and rapid spread of infection in the absence of any intervention. In the event that the introduction of vaccination occurs in the fourth month, it results in a slower and more controlled growth in the number of cases accumulated after the start of vaccination. This indicates that early vaccination has a significant impact in reducing the spread of infection and making the system more stable. When vaccination started in the sixth month, it also shows a reduction in cumulative case growth, although not as effective as vaccination started in the fourth month. This suggests that although late vaccination is still beneficial, its impact is less compared to early vaccination. The case with vaccination started in the 12th month shows the least impact in reducing the growth of cumulative cases. The infection continues to spread more widely before vaccination begins to have a significant effect, resulting in a higher number of cumulative cases compared to the early vaccination scenarios.

The model shows that the stability of the epidemiological system is directly related to when vaccination begins. In the absence of vaccination, the system remains unstable, with a continuous increase in the number of cases. Early introduction of vaccination, such as in the fourth or sixth month, promotes faster stabilization of the system, resulting in a significant reduction in the accumulated cases. However, late vaccination, although still effective, is associated with greater initial instability and a higher number of accumulated cases before stability is achieved.

## Final Remarks

In this study, we conducted a comparative analysis of three mathematical models for directly transmitted epidemics, highlighting the inclusion of social behaviour and vaccination as central elements in the dynamics of infectious disease spread. The simulations demonstrated that the adoption of social distancing strategies effectively reduces the number of active cases temporarily and delays the spread of the epidemic. However, relaxation of these strategies often leads to new outbreaks, emphasizing the need for continuous and well-structured planning of containment measures.

The results also show that early vaccination plays a crucial role in stabilizing the epidemiological system. Early vaccination not only significantly reduces the number of cumulative cases, but also ensures greater control over the epidemic dynamics. In contrast, delayed vaccination, while still beneficial, has a reduced impact and is associated with greater initial instability, underscoring the importance of timely implementation of interventions.

Furthermore, modelling heterogeneous social behaviours proved essential for a better understanding of the interaction between different population groups and their transmission dynamics. Dividing the population into subgroups with varying levels of adherence to mitigation strategies allowed a more detailed analysis of the effects of social distancing and vaccination in realistic scenarios.

The findings of this study reinforce the importance of incorporating behavioural aspects and non-pharmaceutical interventions into epidemiological models, contributing to a more accurate understanding of epidemic dynamics. In addition, the comparative analysis of the proposed models provides valuable information for the formulation of more effective public policies to control infectious diseases.

Future work could explore additional factors, such as the impact of awareness campaigns, variations in vaccine efficacy, and changes in social behaviour over time. It would also be worthwhile to investigate the interaction between multiple simultaneous interventions, further expanding the applicability of the proposed models.

## Data Availability

The data is obtained from the papers cited in the manuscript.

## Notes

### Competing Interest Statement

The authors have declared no competing interest.

### Funding Statement

The author(s) received no specific funding for this work.

### Author Declarations

No ethical approval it is necessary to this work.

